# The Target48 Neurodegeneration Panel: A Novel Tool for Profiling Protein Signatures in Neurodegenerative Disorders

**DOI:** 10.64898/2026.06.15.26355662

**Authors:** Yanaika S. Hok-A-Hin, Lisa Vermunt, Natascha de Jong, Marta del Campo Milan, Everard Vijverberg, Afina W. Lemstra, Yolande A.L. Pijnenburg, Argonde C. Van Harten, Charlotte E. Teunissen

## Abstract

**Introduction:** Novel tools for absolute quantification of established and emerging fluid neuro-biomarkers are required to advance diagnostic studies and improve biological insights.

**Methods:** We conducted an extensive analytical and clinical validation of the Olink™ Target 48 Neurodegeneration panel (T48 Neuropanel) in 352 paired CSF and plasma samples from cognitively unimpaired controls (CU), Alzheimer’s dementia (AD), frontotemporal dementia (FTD), and dementia with Lewy bodies (DLB), n=44 per group. Comparisons with benchmark assays were performed.

**Results:** Good detectability (CSF: 31 out of 42 assays; plasma: 38 out of 42 assays) and technical performance was observed. Benchmark assays showed good correlations, supporting transformation formula’s. Next to emerging biomarkers (MMP10, ITGB2), discriminative performance was excellent in AD: CSF pTau217: AUC=1; FTD: plasma NfL: AUC=0.952; and DLB: CSF DDC: AUC=0.901.

**Discussion:** This analytical and clinical validation of the T48 Neuropanel highlights initial cut-offs and emerging biomarkers to aid clinical studies for the diagnosis, prognosis, and monitoring of neurodegenerative diseases.

**Highlights:**

– The T48 Neuropanel shows robust analytical performance, with high detectability across both plasma and CSF matrices.
– The T48 Neuropanel validates established (i.e., pTau217, Aβ42, NfL, and GFAP) and emerging biomarkers (i.e., DDC, MMP10, ITGB2, ITGAM, NPTX2, NPTXR, SMOC1, sTREM1, and sTREM2) in CSF and plasma.
– CSF NfL, GFAP, ITGB2, and ITGAM and plasma GFAP were dysregulated across AD, FTD, and DLB dementias.
– The multiplex design of the T48 Neuropanel enables rich biological interpretation by simultaneously quantifying established and emerging neurodegeneration biomarkers. Importantly, the inclusion of absolute quantification facilitates the establishment of cut-offs, supporting its potential for clinical translation.

## Introduction

Neurodegenerative dementias, including Alzheimer’s disease (AD), frontotemporal dementia (FTD), and dementia with Lewy bodies (DLB), affect millions of people globally [1, 2]. These conditions are defined by the accumulation of protein aggregates in the brain: AD by amyloid-β plaques and tau tangles, FTD by tau or TDP-43 aggregates, and DLB by α-synuclein aggregates [3–5]. Quantification of brain aggregate pathology and all aspects of neurodegeneration in its full complexity during life remains a challenge. For AD, advanced cerebrospinal fluid (CSF) and blood-based biomarkers (e.g., pTau217) are incorporated into the Alzheimer’s Association criteria for identification of the core AD pathologies [6]. However, for other neurodegenerative dementias, fluid biomarkers for clinical use are not yet available. For FTD, only biofluid measurements of NfL are incorporated into the clinical guidelines to help distinguish FTD from primary psychiatric disorders [7]. In DLB, CSF α-synuclein SAA (αSyn-SAA) has undergone extensive validation and can reliably detect α-synuclein aggregates, and is therefore included in the neuronal α-synuclein disease–integrated staging system research framework [8, 9]. In addition, recent studies highlight dopa decarboxylase (DDC) in CSF as a potential biomarker of Lewy body disorders, likely reflecting dopaminergic dysfunction [10–14]. In view of the advancement of disease-modifying therapies, the need for early, accurate, and pathology-specific diagnosis is becoming increasingly critical. Furthermore, there is a need for fluid biomarkers that can support clinical trials by capturing diverse biological treatment effects [15]. To comprehensively assess similarities and differences between neurodegenerative dementias, the use of protein assay panels that capture diverse underlying brain pathology and biology prove valuable. Recent studies therefore support the added value of multiplex biomarker panels for detecting distinct AD, FTD, and DLB pathogenesis. Such biomarker panels provide added value by capturing the complexity of disease processes in dementias [10, 16–20]. Furthermore, as biomarker profiles are known to shift across disease stages, multiplex panels may have particularly valuable in tracking these dynamic changes along the disease continuum [21, 22].

The Target 48 Neurodegeneration Panel (T48 Neuropanel) is a novel multiplex test based on the proximity extension assay (PEA) technology. This biomarker panel simultaneously quantifies 42 proteins from a low sample volume (1 µL). It is designed to cover a broad range of biological mechanisms often dysregulated in neurodegenerative dementias, such as synaptic (dys)function, neuroinflammation, myelin integrity, axon-glia interactions, neuroprotection, oxidative stress, and vascular functioning. One key advantage of this panel is that it measures proteins in absolute concentrations. This facilitates the development of robust cut-offs and enables external model validation and prospective validation studies, ultimately supporting more accurate biological interpretation and clinical translation. Here, we present the first assessment of this panel in paired plasma and CSF samples from cognitively unimpaired individuals and patients with AD, FTD, and DLB. The aim was to conduct an analytical and clinical validation of the T48 Neuropanel. Analytical performance was assessed by evaluating cross-reactivity between the assays in the T48 Neuropanel, the detectability in CSF and in the optimized matrix (plasma), and by comparing T48 Neuropanel measurements with results from orthogonal methods. The clinical validation included comparison of the T48 Neuropanel protein levels in AD, FTD, and DLB to the levels in controls, assessment of the discriminative power through ROC curve analysis, with the Youden cut-offs. Lastly, we assessed which biomarker abnormalities are shared across these neurodegenerative dementias.

## Methods

### Human CSF and plasma samples

Three hundred fifty-two paired CSF and plasma samples from cognitively unimpaired controls (n = 44) and individuals diagnosed with AD (n = 44), FTD (n = 44), and DLB dementia (n = 44) from the Amsterdam Dementia Cohort (ADC) were included [23, 24]. Among these, 8 samples were unpaired (6 AD, 2 FTD, and 1 DLB) because either the CSF or plasma sample was unavailable. Cohort characteristics are shown for the paired samples in Table 1. All participants underwent standard neurological screening and cognitive testing in the ADC. Clinical diagnosis were made in a multidisciplinary consensus meeting based on the diagnostic criteria for each type of dementia [6, 25–28]. CSF was collected and stored according to consensus biobanking guidelines [29]. EDTA plasma samples were obtained through venipuncture, centrifuged at 1800 x g for 10 minutes within 2 hours of collection, and stored at –80°C until analysis, following SOP guidelines [30]. Concentrations of the core AD CSF biomarkers were used to support the diagnosis of AD and to determine AD copathology in DLB patients, measured by commercially available ELISA kits (Innotest Aβ(1–42) and Innotest hTAUAg assays, Fujirebio, Ghent, Belgium) or by the Elecsys platform (Elecsys β-Amyloid (1-42) CSF II and Elecsys Phospho-Tau (181P) CSF assay, Roche Diagnostics, Gmbh, Almere, The Netherlands). A positive AD profile was defined as a tTau/Aβ42 ratio > 0.46 (ELISA) or pTau181/Aβ42 > 0.020 (Elecsys)[31, 32]. CSF Aβ42 concentrations measured by ELISA were corrected for the drift that occurred over the years, as described previously [33]. The individuals with FTD included in this study were clinically diagnosed with behavioral variant FTD [34]. Individuals with subjective cognitive decline who had normal clinical and cognitive test results and a negative core AD CSF biomarkers (ELISA: tTau/Aβ42 ratio < 0.46; Elecsys: pTau181/Aβ42 ratio < 0.020) were included as controls [31, 32]. Informed consent was obtained from all participants or their authorized representatives, in accordance with the ethical approval of the Amsterdam UMC IRB and in agreement with the Helsinki Declaration of 1975.

**Table 1.** Cohort characteristics.

|  |  | **Controls** | **AD** | **FTD** | **DLB** |
| --- | --- | --- | --- | --- | --- |
| n |  | 44 | 38 | 42 | 43 |
| Sex, n (%) | F | 17 (39) | 20 (53) | 18 (43) | 3 (7) |
|  | M | 27 (61) | 18 (47) | 24 (57) | 40 (93) |
| Age (years) |  | 53 (49 - 57)<sup>a,b,c</sup> | 66 (59 - 72)<sup>d</sup> | 62 (57 - 67)<sup>b,d</sup> | 69 (64 - 72)<sup>c,d</sup> |
| AD CSF Biomarker Profile, n (%) | Negative | 44 (100) | 0 (0) | 36 (95) | 27 (64) |
|  | Positive | 0 (0) | 38 (100) | 2 (5) | 15 (36) |
| MMSE scores |  | 29 (27 - 29)<sup>a,b,c</sup> | 19 (15 - 24)<sup>b,c,d</sup> | 25 (22 - 28)<sup>a,d</sup> | 24 (21 - 26)<sup>c,d</sup> |

Cohort characteristics for the samples for which both CSF and plasma is available is shown.
Continuous data is represented as median ± interquartile range and dichotomous data as the number of cases with a percentage of the total (%).
Differences between groups were determined using the Kruskal Wallis test with Bonferroni correction or Chi-squared test.
a, p < 0.05 compared to AD
b, p < 0.05 compared to DLB
c, p < 0.05 compared to FTD
d, p < 0.05 compared to Controls
Abbreviations: AD, Alzheimer's disease; FTD, frontotemporal dementia; DLB, dementia with Lewy bodies; MMSE, mini-mental state examination; F, female, M, male, CSF, cerebrospinal fluid.

### Protein analysis

#### Target 48 Neurodegeneration Panel

Forty-two proteins were simultaneously measured using the multiplex T48 Neuropanel (kit lot: #LC00406, Olink Proteomics, Uppsala, Sweden). Measurements were performed on the Q100 instrument (Olink Proteomics) in the Neurochemistry Laboratory (Amsterdam UMC, Amsterdam, The Netherlands). All experimental procedures were performed by technicians blinded to the diagnostic information of the samples. CSF and plasma samples were randomized based on clinical diagnosis across multiple plates, separated by matrix. Each plate additionally included three quality controls (QCs), two negative controls, and three single-point calibrators. Precision (intra- and inter-assay coefficients of variation; CV) of the assays was calculated using three QC samples provided by the manufacturer. For each protein, a 24-point standard curve was established by the manufacturer using four-parameter logistic (4PL) curve fitting. The median values of the single-point calibrators were used to adjust to the 24-point predefined calibration curve. The lower and upper limits of quantification (LLOQ and ULOQ) were defined by the manufacturer during panel development. Assays were excluded from further analysis if protein concentrations were below the LLOQ or above the ULOQ in more than 15% of the samples. Protein concentrations were reported as absolute concentrations (pg/mL).

#### Specificity test

To determine cross-talk between the 42 proteins within the T48 Neuropanel, specificity tests were performed as part of standard panel development by Olink Proteomics and data was kindly shared with the authors for use. Recombinant antigens were serially diluted to achieve the desired target concentrations (detailed in Supplementary Table 1). The target concentration for each recombinant antigen was determined based on the signal intensity observed in EDTA plasma samples analyzed in-house for each assay. All 42 antigens were assayed in duplicate, and a pooled mixture of all 42 antigens was included as a positive control in each assay. The contribution of nonspecific signal was calculated by dividing the nonspecific signal by the true signal.

#### NULISA CNS Panel

The same CSF and plasma samples were analyzed using the NULISAseq CNS Panel (NULISA_120-CNS Panel_), targeting 120 proteins associated with a broad spectrum of neurodegenerative disorders, as part of our previous work (*B. Ariöz & Y.S. Hok-A-Hin et al., under review)*. All samples were randomized prior to analysis. Next-generation sequencing data were processed using the NULISAseq algorithm (Alamar Biosciences), and intra-plate and inter-plate normalization were performed as described previously (*B. Ariöz & Y.S. Hok-A-Hin et al., under review*). The CV% were based on 2 sample controls included on each plate and were < 15%. The LOD for each assay was determined as 3 standard deviations above the negative control. Assays were excluded from further analysis if protein levels were below the LOD in more than 15% of the samples. Interpolated control-normalized counts were log_2_-transformed and referred to as NULISA Protein Quantification (NPQ) units.

#### Simoa Neurology 4-PLEX

Concentrations of plasma NfL, GFAP, Aβ42, and Aβ40 (Simoa Neurology 4-PLEX E Advantage kit; Quanterix Corp, kit lot: #503143) were previously determined in a subset of the cohort (n = 67) on the Simoa HD-X Analyzer (Quanterix, Billerica, USA) [35]. Plasma samples were tested in single measurements with on-board automated sample dilution, according to the manufacturer’s instructions. Two quality control samples were included in all Simoa runs, showing good inter-assay coefficients of variation < 15% for all biomarkers.

### Statistical analysis

All statistical analyses were performed in Rstudio (version 2023.12.1). Patient demographics were tested using the Kruskal-Wallis test followed by Bonferroni correction for continuous variables or Pearson’s chi-square test for categorical variables. Absolute values measured with the T48 Neuropanel were Log_2_-transformed for all analysis where normal distribution was required.

The T48 Neuropanel (Log_2_) values were compared to protein levels measured by NULISA for the overlapping proteins (n = 16, Supplementary Table 2) using Pearson correlation analysis. Spearman correlation analysis was performed to directly compare between protein concentrations measured by the T48 Neuropanel (pg/mL) and Elecsys (i.e., Aβ42 assay); and between T48 Neuropanel (pg/mL) and Simoa (NfL, GFAP, Aβ42, and Aβ40 assays). In addition, Passing–Bablok regression and Bland–Altman analyses were performed to assess platform agreement for the T48 Neuropanel with Elecsys and Simoa assays.

Differences between groups in CSF and plasma log_2_-transformed protein levels were assessed using ANCOVA, corrected for age and sex. The p-values were FDR-corrected for the number of proteins included in the analyses. For the significant differentially regulated proteins, post-hoc analyses included the comparison of each dementia group to the control group, corrected for the total number of group comparisons using the Bonferroni method. Furthermore, to test the discriminative performance, receiver operating characteristics (ROC) analysis were performed using the raw biomarker values. Optimal biomarker cut-offs (pg/mL) were determined based on Youden’s Index for each comparison (AD vs controls, FTD vs controls, and DLB vs controls). P < 0.05 was considered significant.

Spearman correlation analysis was used to assess the correlation for each protein concentration between CSF and plasma matrix. Correlations coefficients Spearman’s *r* or Spearman’s *ρ* < 0.3 are considered weak, between 0.3–0.5 moderate and > 0.5 are considered strong.

## Results

### The T48 Neuropanel shows good detectability and analytical performance in CSF and plasma

We first determined the analytical performance of the novel T48 Neuropanel. Specificity testing revealed that, among the 42 assays, cross-reactivity was observed only for the OMG assay in plasma, which showed a 10% increase in signal when combined with the ITGB2 antigen (Supplementary Figure 1).

Next, the detectability of the assays in CSF and plasma in our cohort was assessed. The results show that 31 proteins (74%) were successfully detected in CSF (Supplementary Figure 2). Eight assays had results below the LLOQ (i.e., LRRK2, GDNF, WWOX, FOXO3, NGF, EIF2AK2, TP53, and SNCAIP) and three assays had results above the ULOQ (i.e., OMG, MOG, and NPTXR) in at least 26 out of 176 samples (15%). The results show that 38 proteins (90%) were successfully detected in plasma (Supplementary Figure 3). Four assays had results below the LLOQ (i.e., S100B, LRRK2, NLGN1, and SNCAIP) in at least 26 out of 176 samples (15%).

The precision of the T48 Neuropanel was good, with a median inter-assay CV of 7.4% and an median intra-assay CV of 6.5% measured in the quality control samples (n = 30). Additional exploration of the 42 individual assays revealed that all assays showed good inter- and intra-assay CVs (all < 15%), indicating reproducible analytical performance across runs for all assays (Supplementary Table 3).

### Cross-platform comparison shows moderate to strong correlations across multiple methods

We next assessed correlations between the T48 Neuropanel and other immune-assay based platforms. A strong correlation between concentrations obtained with the CSF Elecsys Aβ42 and the T48 Aβ42 assays was observed (n = 28, *ρ* = 0.91, p < 0.001; Figure 1 A-C). Passing-Bablok analysis revealed no platform agreement with higher levels measured by the T48 Neuropanel (Supplementary Figure 4A). After transforming the T48 Neuropanel values to Elecsys using the Passing-Bablok transformation formula, values were comparable between methods (Supplementary Figure 4B). For the comparison in CSF with NULISA_(120-CNS panel),_ 14 of the 16 overlapping assays had a strong correlation with the T48 Neuropanel (*r* between 0.58 – 0.95, all p < 0.001; Figure 1A), of which pTau217 and NfL showed the strongest correlation (both: *r* = 0.95, p < 0.001). Passing-Bablok analysis was not performed because the NULISA_(120-CNS Panel)_ does not generate absolute quantification of the protein measurements.

**Figure 1.**
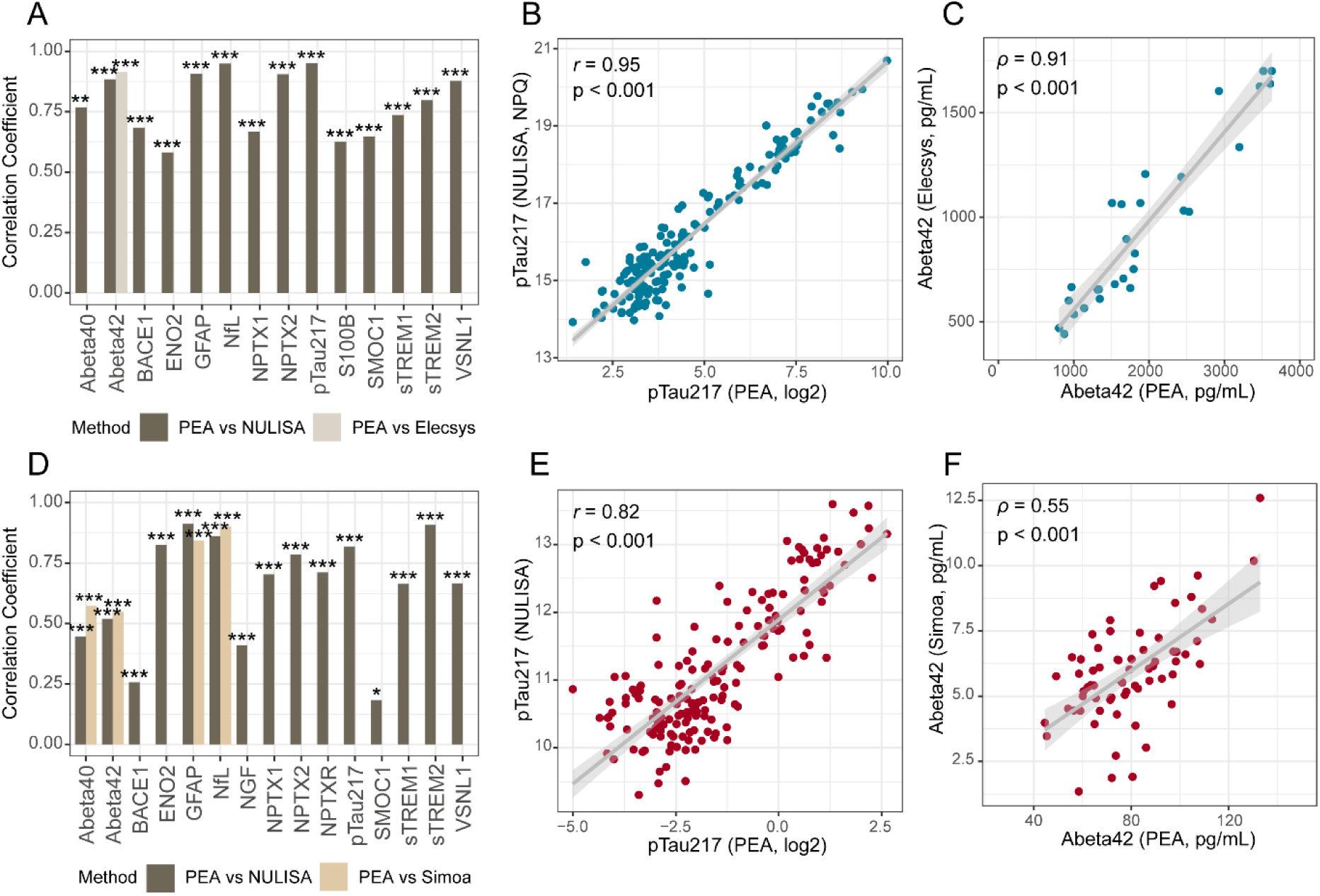
Cross-platform comparison of the T48 Neuropanel in CSF and plasma. Overall, moderate to strong correlations were observed in CSF across different immune-based assays. Bars represent the Pearson’s *r* or Spearman’s *ρ* correlation coefficient for each assay comparison in CSF (A) and plasma (D). Strong correlations were observed in CSF for pTau217 (B) and Aβ42 (C) when comparing the T48 Neurodegeneration panel with NULISA and Elecsys, respectively. In plasma, correlations between immune-based assays were generally moderate to strong, except for BACE1 and SMOC1 (D). Strong correlations between T48-PEA and NULISA were observed for plasma pTau217 (E), whereas plasma Aβ42 showed moderate correlations between T48-PEA and Simoa (F). Abbreviations: PEA, proximity extension assay; NPQ, NULISA Protein Quantification. *indicates < 0.05, **indicates p < 0.01, ***indicates p < 0.001.

In plasma, strong correlations were observed between the T48 Neuropanel and Simoa NfL (*ρ* = 0.90, p < 0.001), GFAP (*ρ* = 0.84, p < 0.001), Aβ42 (*ρ* = 0.55, p < 0.001), and Aβ40 (*ρ* = 0.57, p < 0.001) concentrations (Figure 1D-F). None of these assays showed platform agreement, and thus Passing-Bablok transformation formulas were calculated (Supplementary Figure 5). After transforming the T48 Neuropanel values to the Simoa scale, no significant bias between methods was observed, indicating that the protein values between platforms are on the same scale (Supplementary Figure 5). Visual inspection of the Bland-Altman plots of Aβ42 and Aβ40 showed a minor trend towards increasing underestimation of the biomarker values at higher concentrations. This suggests that the biomarker values are not fully aligned across the full analytical range. Furthermore, results for 11 assays showed a strong correlation between the T48 Neuropanel and the NULISA_(120-CNS Panel)_ platforms (*r* between 0.52 – 0.91, all p < 0.001; Figure 1D). Noteworthy, 3 assays had a weak to moderate correlation in plasma between these platforms (SMOC1, BACE, and NGF; *r* between 0.18 – 0.45, all p < 0.001; Figure 1D).

Overall, the T48 Neuropanel results correlated well with other immunoassay-based methods, particularly for the established biomarkers NfL, GFAP, Aβ42, Aβ40, and pTau217, which showed slightly stronger correlations in CSF than in plasma. Passing-Bablok formulas to convert NfL, GFAP, Aβ42, and Aβ40 on the T48 Neuropanel aligned well with Elecsys and Simoa measurements.

### CSF shows strong protein dysregulations and discriminative performance across neurodegenerative dementias

We next determined the protein profiles of AD, FTD, and DLB dementias compared to controls. Concentrations of 18 CSF proteins (17 increased and 1 decreased) were dysregulated in AD. The 3 CSF proteins with the largest fold changes were pTau217, Aβ42, and ITGB2 (Figure 2A). These proteins showed excellent discriminative performance compared to controls (pTau217: AUC = 1.00, 95%CI: 1.00-1.00; Aβ42: AUC = 0.98, 95%CI: 0.97-1.00; ITGB2: AUC = 0.97, 95%CI: 0.94-1.00; Figure 2D). We additionally determined their cut-offs using Youden’s Index (Table 2).

**Figure 2.**
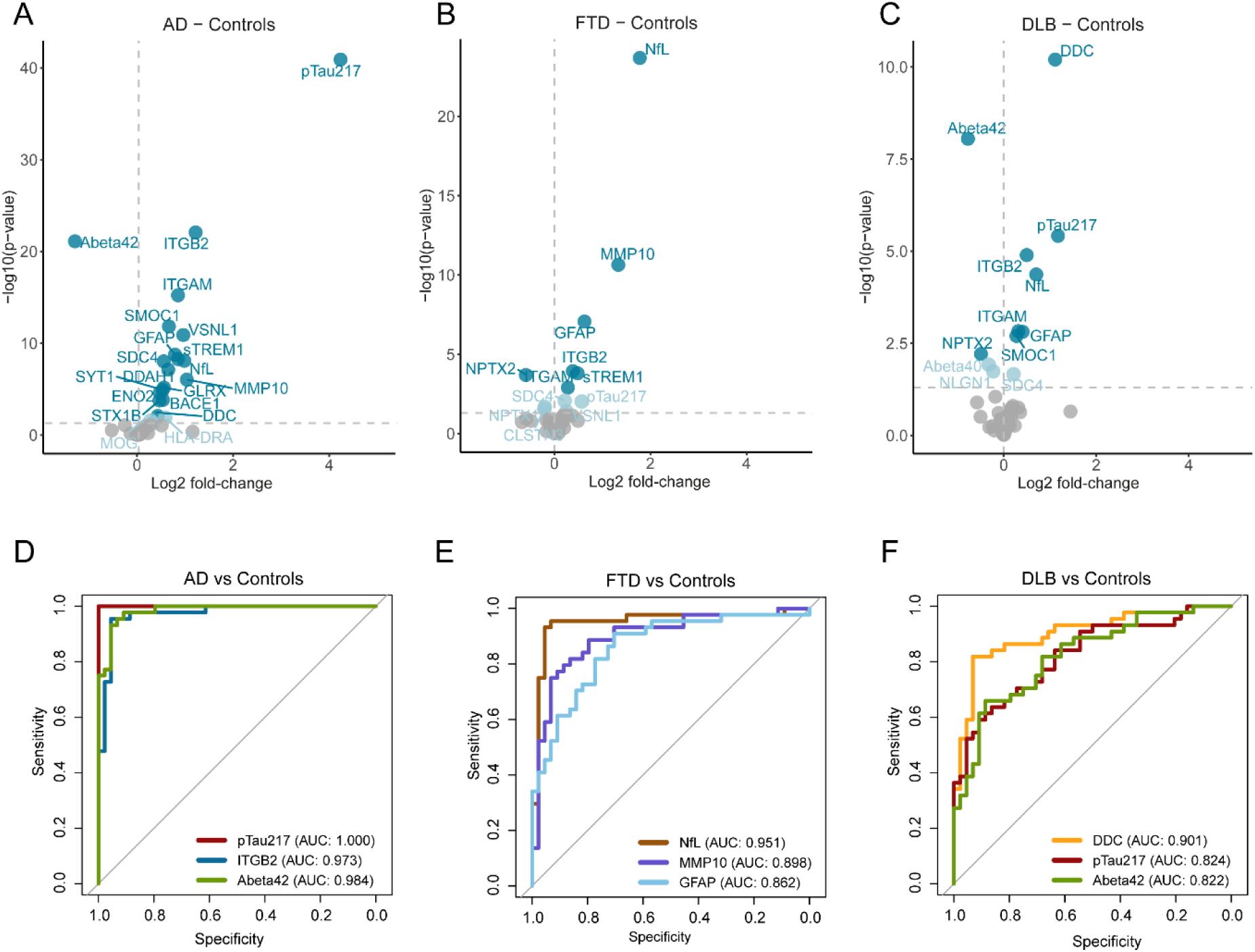
CSF protein dysregulation and discriminative performance. Volcano plots show the differential protein abundance in plasma between AD (A), FTD (B), and DLB (C) compared to controls. The differences (log2 fold-change) are plotted against the q-value (-log10 transformed). FDR-corrected CSF proteins that are significantly dysregulated are shown in light-blue, FDR-corrected CSF proteins after correction for multiple testing by Bonferroni are shown in blue. Each dot represents a protein and the horizontal line shows the threshold for significance. ROC curve analysis was performed to determine the discriminative performance for the top 3 proteins that had the strongest fold-change in CSF to discriminate (D) AD from controls (pTau217: AUC = 1, 95%CI: 1-1; Aβ42: AUC = 0.984, 95%CI: 0.965 −1; and ITGB2: AUC = 0.973, 95%CI: 0.941-1), (E) FTD from controls (NfL: AUC = 0.951, 95%CI: 0.890-1; MMP10: AUC = 0.898, 95%CI: 0.829-0.966; and GFAP: AUC = 0.862, 95%CI: 0.784-0.939), and (F) DLB from controls (DDC: AUC = 0.901, 95%CI: 0.835-0.967; pTau217: AUC = 0.824, 95%CI: 0.738-0.910; and Aβ42: AUC = 0.822, 95%CI: 0.735-0.908). Abbreviations: AD, Alzheimer’s disease; FTD, frontotemporal dementia; DLB, dementia with Lewy bodies; MMSE, mini-mental state examination

**Table 2.** Optimal biomarker cut-offs with corresponding performance parameters.

| **Biomarker** | **Comparison** | **Matrix** | **Cut-off (pg/mL)** | **Sensitivity (%)** | **Specificity (%)** |
| --- | --- | --- | --- | --- | --- |
| pTau217 | AD vs Controls | CSF | 37.61 | 100% | 100% |
| ITGB2 | AD vs Controls | CSF | 696.89 | 95% | 95% |
| Abeta42 | AD vs Controls | CSF | 1769.38 | 98% | 91% |
| NfL | FTD vs Controls | CSF | 11663.90 | 95% | 93% |
| MMP10 | FTD vs Controls | CSF | 12.70 | 82% | 86% |
| GFAP | FTD vs Controls | CSF | 968.79 | 91% | 70% |
| DDC | DLB vs Controls | CSF | 22.50 | 82% | 93% |
| Abeta42 | DLB vs Controls | CSF | 1911.13 | 66% | 89% |
| pTau217 | DLB vs Controls | CSF | 14.97 | 64% | 86% |
| pTau217 | AD vs Controls | Plasma | 0.49 | 95% | 100% |
| GFAP | AD vs Controls | Plasma | 20.84 | 84% | 93% |
| NPTXR | AD vs Controls | Plasma | 1421.49 | 93% | 34% |
| NfL | FTD vs Controls | Plasma | 682.17 | 88% | 95% |
| GFAP | FTD vs Controls | Plasma | 13.85 | 93% | 70% |
| Abeta42 | FTD vs Controls | Plasma | 81.15 | 74% | 57% |
| GFAP | DLB vs Controls | Plasma | 13.30 | 88% | 68% |
| pTau217 | DLB vs Controls | Plasma | 0.42 | 42% | 98% |
| NfL | DLB vs Controls | Plasma | 644.40 | 74% | 91% |

Abbreviations: AD, Alzheimer's disease; FTD, frontotemporal dementia; DLB, dementia with Lewy bodies; CSF, cerebrospinal fluid.

In FTD, 7 CSF proteins (6 increased and 1 decreased) were dysregulated, of which NfL, MMP10, and GFAP concentrations showed the strongest increase compared to controls (Figure 2B). These proteins additionally showed good to excellent performance in differentiating FTD from controls (NfL: AUC = 0.95, 95%CI: 0.89-1.00; MMP10: AUC = 0.90, 95%CI: 0.83-0.97; GFAP: AUC = 0.86, 95%CI: 0.78-0.94; Figure 2E). NfL, MMP10 and GFAP cut-offs were determined to classify FTD from controls (Table 2).

Concentrations of 9 CSF proteins (7 increased and 2 decreased) were dysregulated in DLB. The largest protein changes were observed for DDC, Aβ42, and pTau217 proteins (Figure 2C), which additionally showed good to excellent discriminative performance (DDC: AUC = 0.90, 95%CI: 0.84-0.97; Aβ42: AUC = 0.82, 95%CI: 0.74-0.91; pTau217: AUC = 0.82, 95%CI: 0.74-0.91; Figure 2F). Their cut-offs were determined to classify DLB from controls (Table 2).

Notably, 4 CSF proteins (i.e., GFAP, NfL, ITGB2, and ITGAM) were significantly altered across AD, FTD, and DLB dementia (Supplementary Figure 6A). In addition, 8 CSF proteins were uniquely dysregulated in AD (i.e., VSNL1, SDC4, DDAH1, SYT1, GLRX, ENO2, BACE1, and STX1B), whereas no uniquely dysregulated CSF proteins were identified in FTD or DLB (Supplementary Figure 6A).

### Plasma protein dysregulation is limited across neurodegenerative dementias, with moderate to strong discriminative performance

We next evaluated protein changes in plasma for AD, FTD, and DLB dementias. Concentrations of 2 plasma proteins were increased (i.e., pTau217 and GFAP) and of 1 protein were decreased (NPTXR) in AD compared to controls (Figure 3A). pTau217 and GFAP showed excellent performance to discriminate AD from controls, while the performance of NPTXR was poor (pTau217: AUC = 0.97, 95%CI: 0.92-1.00; GFAP: AUC = 0.94, 95%CI: 0.90-0.99; NPTXR: AUC = 0.60, 95%CI: 0.47-0.72; Figure 3D). In addition, their cut-offs were determined to classify AD from controls using Youden’s Index (Table 2).

**Figure 3.**
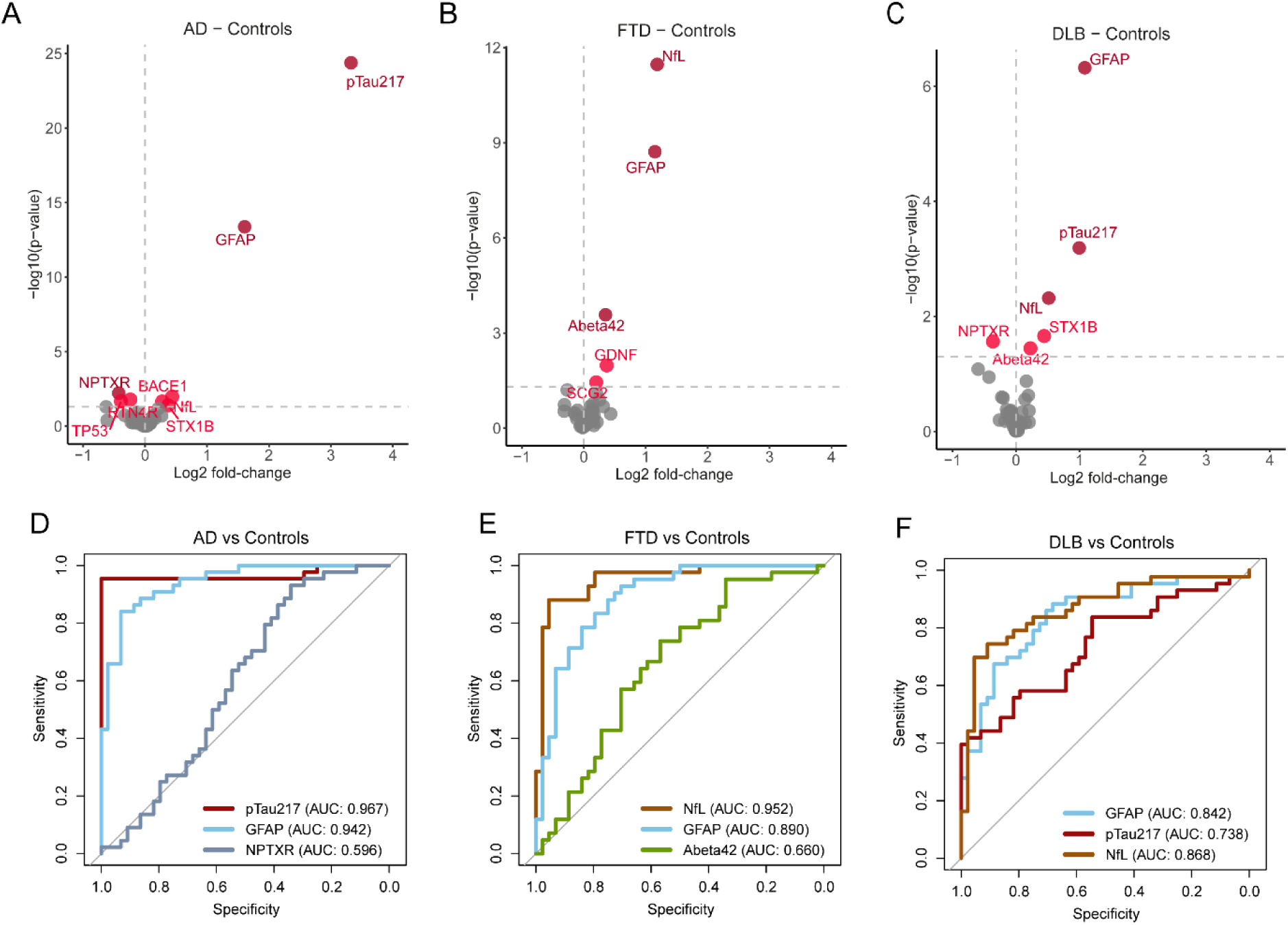
Plasma protein dysregulation and discriminative performance. Volcano plots show the differential protein abundance in plasma between AD (A), FTD (B), and DLB (C) compared to controls. The differences (Log2 fold-change) are plotted against the corrected p-value (-log10 transformed). FDR-corrected CSF proteins that are significantly dysregulated are shown in red, FDR-corrected CSF proteins after correction for multiple testing by Bonferroni are shown in dark red. Each dot represents a protein and the horizontal line shows the threshold for significance. ROC curve analysis was performed to determine the discriminative performance for the top 3 proteins that had the strongest fold-change in plasma to discriminate (D) AD from controls (pTau217: AUC = 0.967, 95%CI: 0.921-1; GFAP: AUC = 0.942, 95%CI: 0.897-0.987; and NPTXR: AUC = 0.596, 95%CI: 0.474-0.718), (E) FTD from controls (NfL: AUC = 0.952, 95%CI: 0.908-0.997; GFAP: AUC = 0.890, 95%CI: 0.821-0.959; and Aβ42: AUC = 0.660, 95%CI: 0.543-0.776) and (F) DLB from controls (GFAP: AUC = 0.842, 95%CI: 0.758-0.926; pTau217: AUC = 0.738, 95%CI: 0.633-0.844; and NfL: AUC = 0.868, 95%CI: 0.789-0.947).Abbreviations: AD, Alzheimer’s disease; FTD, frontotemporal dementia; DLB, dementia with Lewy bodies; MMSE, mini-mental state examination

Three proteins were elevated in plasma of FTD patients compared to controls (i.e., NfL, GFAP and Aβ42; Figure 3B). ROC curve analysis showed excellent discriminative performance for NfL and GFAP, whereas the performance of Aβ42 was relatively poor (NfL: AUC = 0.95, 95%CI: 0.91-0.99; GFAP: AUC = 0.89, 95%CI: 0.82-0.96; Aβ42: AUC = 0.66, 95%CI: 0.54-0.78; Figure 3E). Cut-offs for these plasma proteins were additionally determined that classify FTD from controls (Table 2).

Concentrations of three plasma proteins were increased in DLB compared to controls (i.e., GFAP, pTau217, and NfL; Figure 3C). These proteins showed moderate to good performance for discriminating DLB from controls (GFAP: AUC = 0.84, 95%CI: 0.76-0.93; pTau217: AUC = 0.74, 95%CI: 0.63-0.84; NfL: AUC = 0.87, 95%CI: 0.79-0.95; Figure 3F). Their cut-offs were determined to classify DLB from controls (Table 2).

Plasma GFAP was the only plasma protein significantly altered across AD, FTD, and DLB (Supplementary Figure 6B). In addition, NPTXR was uniquely dysregulated in AD, and Aβ42 was uniquely dysregulated in FTD, while no proteins were uniquely dysregulated in DLB (Supplementary Figure 6B).

### Moderate to strong protein correlations between CSF and plasma

Comparison of concentrations between CSF and plasma was possible for 30 proteins. Concentrations of 43% of these proteins correlated significantly between CSF and plasma (*ρ* between 0.15 – 0.79). The strongest correlations were observed for NfL (*ρ* = 0.79), pTau217 (*ρ* = 0.71), and AGER (*ρ* = 0.64; Figure 4). Interestingly, many proteins that were not correlated are representing synaptic function and dysfunction (i.e., NPTX2, NPTXR, SYT1, KLK8, STX1B, SCG2, and CLSTN3). Upon stratifying the cohort by diagnosis, some proteins correlated between matrices only in the controls (e.g., VSNL1), whereas others correlated specifically in the dementia groups (e.g., GFAP and sTREM2).

**Figure 4.**
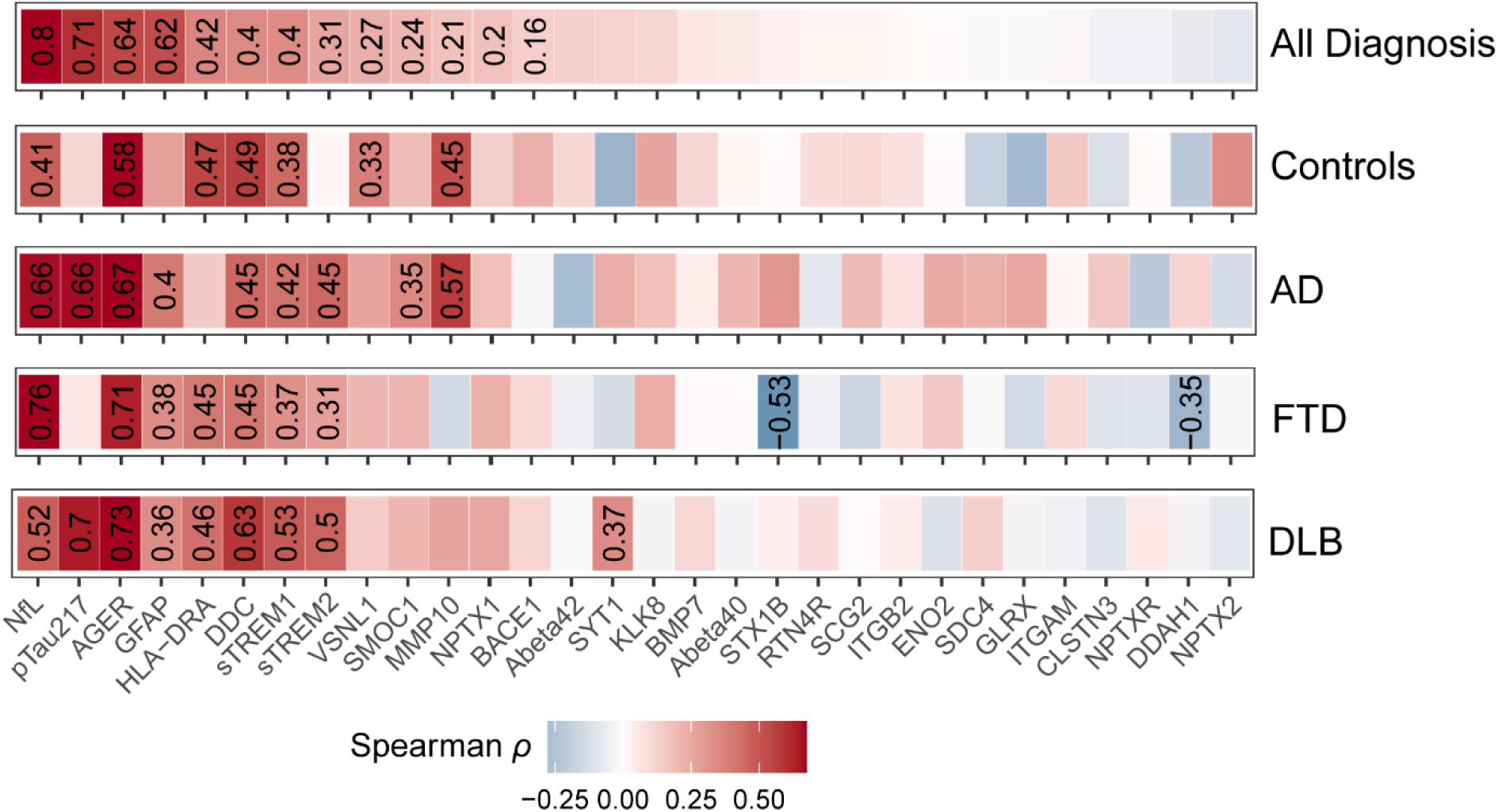
Correlations between CSF and plasma matrix. Correlations between CSF and matrix matric were determined in 30 proteins by Spearman correlation analysis. Heatmap shows the Spearman’s *ρ* correlation coefficient of proteins measured in CSF compared to plasma in the total cohort and stratified for clinical diagnosis. Only for the significant correlations the Spearman’s ρ are indicated with the number, non-significant correlation coefficients are not shown in the heatmap. The red color depicts a positive correlation coefficient, and the blue color depicts a negative correlation coefficient. Abbreviations: AD, Alzheimer’s disease; FTD, frontotemporal dementia; DLB, dementia with Lewy bodies.

## Discussion

In this study, we present the first analytical and clinical evaluation of the T48 Neuropanel in paired CSF and plasma from patients with different neurodegenerative dementias. The T48 Neuropanel shows good detectability (CSF: 74%; plasma: 90% of the 42 assays) and correlates well with other immune-based methods. The panel contains established (i.e., pTau217, Aβ42, NfL, and GFAP) and emerging biomarkers (i.e., ITGB2, MMP10, DDC), which showed good discriminative performance between groups. We additionally provide transformation formulas and clinical cut-offs for use in future studies. Lastly, we established correlations between CSF and plasma and showed the extent to which these correlations depended on the presence of neurodegenerative disease.

Fluid biomarkers have revolutionized the detection, diagnosis, and screening of neurodegenerative dementias, especially for AD [36, 37]. The potential of the T48 Neuropanel lies in its capability to measure the concentrations of 42 proteins in a small volume of sample (1 µL), thereby reducing the time required for analysis and minimizing the amount of sample needed. Considering the novelty of this panel, we first assessed the detectability in CSF and plasma and performed a cross-platform comparison with established immunoassay technologies, namely Elecsys, Simoa, and NULISA methods. Good detectability of the 42 proteins in CSF (76%) and plasma (90%) was observed. Notably, the reduced detectability in CSF relative to plasma can be attributed to the optimization of the assays for plasma samples. In addition, the lower detectability of proteins in CSF results from 3 proteins exceeding the assays upper quantification limits following the dilution required for CSF. LRRK2 and SNCAIP were not detected in either CSF or plasma, which is likely due to the cohort selection, as these markers have been associated with Parkinson’s disease [38, 39]. The data showed an overall good correlation between platforms. Noteworthy, given that absolute quantification is a key advantage of the T48 Neuropanel, platform agreement was assessed to evaluate whether measurements across methods fall within a comparable range. No platform agreement was observed, which is likely due to differences in antibodies and assay setup between these technologies, which is consistent with literature showing similar observations for many plasma-based assays [40–42]. Therefore, the current study additionally provides lot-specific Passing-Bablok formula’s and cohort-specific cut-offs for established biomarkers (i.e., Aβ42, Aβ40, GFAP, and NfL) as a reference for future studies. However, as batch-to-batch variation between kits may influence the transformation formula’s, and cut-offs are inherently cohort-specific, external validation across independent cohorts will be necessary to confirm their generalizability and evaluate cross-laboratory variability.

To further explore disease-specific protein differences and better understand the underlying pathological mechanisms across neurodegenerative disorders, we next assessed the differential protein abundance in AD, FTD, and DLB across matrices. Within CSF, we observed a wide range of protein changes across these neurodegenerative disorders. Many of the strongly dysregulated proteins in CSF are well-established dementia biomarkers (e.g., pTau217, Aβ42, GFAP, and NfL). In addition, emerging CSF biomarkers were also detected (e.g., ITGB2, MMP10, and DDC). ITGB2 forms a heterodimeric receptor with ITGAM and is involved in cell-surface-mediated signaling, with high expression in immune cells including microglia [43]. Multiple proteomic studies have detected increased ITGB2 levels in AD, as well as in earlier clinical stages of AD [19, 21, 43, 44]. MMP10 was recently shown to be increased in patients with FTD, additionally detecting particularly high levels in FTD *MAPT* carriers [20, 45]. Furthermore, DDC showed the strongest fold change in DLB, which corroborates recent work that shows the clinical value of DDC as a diagnostic biomarker for DLB and PD [10, 14]. This, in turn, underscores the value of the T48 Neuropanel as a novel multiplex assay capable of quantitatively detecting DDC, complementing the existing DDC Simoa and DDC Ella assays [14].

Blood-based biomarkers offer a less invasive and more cost-effective approach than CSF, making them highly accessible and effective for tracking disease-related pathological processes. Some of the strong CSF protein changes were also observed in plasma (e.g., pTau217, GFAP, and NfL), in line with observations on other platforms [36]. Furthermore, plasma NPTXR, a marker reflecting synaptic dysfunction, was one of the few plasma proteins that was significantly decreased in AD compared with controls. This is in agreement with literature using other technologies, which showed that higher levels of NPTXR are associated with better cognitive function [46–48]. This highlights the potential of NPTXR as an AD staging biomarker or as a prognostic biomarker.

Due to the paired design of this study, we could evaluate protein levels between CSF and plasma matrices. Protein changes were generally more pronounced in CSF than in plasma, which is likely due to the close association of CSF to the brain pathology. In addition, 43% of the proteins were significantly correlated between CSF and plasma. Interestingly, proteins that did not correlate between matrices are more related to synaptic function and dysfunction, suggesting that peripheral sources may mask CNS-derived changes in blood [49]. When stratifying the analyses by clinical diagnosis, we found that certain proteins correlated between CSF and plasma specifically within the dementia groups (e.g., GFAP and sTREM2). The increased protein levels observed in CSF likely reflect disease-related processes, including enhanced release from reactive astrocytes and increased cleavage of extracellular domains, as described for GFAP and sTREM2, respectively [50, 51]. The positive correlation between CSF and plasma levels suggests that increased CSF concentrations may also contribute to higher plasma concentrations. One potential mechanism is impaired blood–brain barrier integrity in dementia, enabling brain-derived proteins to enter the peripheral circulation [52]. Together, these findings reiterate that CSF remains an important matrix for studying brain diseases.

This study is not without limitations. Specifically, not all proteins from the T48 Neuropanel could be compared with those assessed on other platforms (e.g., pTau217), highlighting the need for future comparative analyses. In addition, other biological matrices such as serum or urine were not evaluated, however, in biomarker studies of neurodegenerative dementias, biobanked samples typically include CSF and plasma. Furthermore, comorbidities such as BMI and chronic kidney disease could influence plasma biomarker levels in clinical disease groups [53, 54]. However, such analyses were outside of the scope of the current study. Lastly, some of our analysis may be limited by the relatively small sample sizes within the diagnostic groups, underscoring the need for validation in larger, independent cohorts. One of the strengths of this study is the workflow we applied. We comprehensively assessed the analytical performance of the T48 Neuropanel and calculated cross-platform transformation formulas. In addition, the paired design of this study allowed us to simultaneously assess protein changes in both CSF and plasma, thereby testing both well-characterized and emerging fluid biomarkers.

### Conclusion

The T48 Neuropanel represents a novel tool for gaining biological insights into various neurodegenerative diseases. Its multiplex design, combined with absolute protein quantification of established and emerging CSF (Aβ42, pTau217, GFAP, NfL, ITGB2, MMP10, and DDC) and plasma biomarkers (Aβ42, pTau217, NfL, GFAP, and NPTXR), enables a rich biological interpretation and supports its potential for clinical translation. We have performed analytical and clinical evaluation of the T48 Neuropanel, additionally providing transformation formulas and clinical cut-offs for future studies.

## Data availability

The data generated in this study is available from the authors on reasonable request.

## Author contributions

YH, LV, and CT conceived and designed the study. YH performed the statistical analysis. YH, LV, MC, EV, AL, AH, YP, and CT recruited participants and collected clinical data and samples. NJ performed laboratory experiments. YH, LV, and CT drafted the manuscript. All authors contributed to the revision and editing of the manuscript.

