## Supplemental Data for "The Target48 Neurodegeneration Panel: A Novel Tool for Profiling Protein Signatures in Neurodegenerative Disorders"

Supplementary Table 1. Protein concentrations used for the specificity tests

| **Index** | **Antigen** | **Uniprot** | **Desired protein concentration for test (ng/mL)** |
|---|---|---|---|
| 1 | VSNL1 | P62760 | 0.042 |
| 2 | FOXO3 | O43524 | 0.105 |
| 3 | sTREM2 | Q9NZC2 | 25.60 |
| 4 | BMP7 | P18075 | 10.24 |
| 5 | SDC4 | P31431 | 10.24 |
| 6 | Abeta40 | P05067 | 0.262 |
| 7 | Abeta42 | P05067 | 0.105 |
| 8 | OMG | P23515 | 0.262 |
| 9 | sTREM1 | Q9NP99 | 0.262 |
| 10 | DDC | P20711 | 10.24 |
| 11 | NPTX1 | Q15818 | 10.24 |
| 12 | NfL | P07196 | 1.638 |
| 13 | pTau217 | P10636 | 0.003 |
| 14 | NGF | P01138 | 0.007 |
| 15 | STX1B | P61266 | 0.105 |
| 16 | S100B | P04271 | 0.017 |
| 17 | ITGAM | P11215 | 10.24 |
| 18 | ITGB2 | P05107 | 64.00 |
| 19 | HLA-DRA | P01903 | 0.105 |
| 20 | GDNF | P39905 | 0.001 |
| 21 | GFAP | P14136 | 0.042 |
| 22 | NPTXR | O95502 | 4.096 |
| 23 | MMP10 | P09238 | 0.655 |
| 24 | SMOC1 | Q9H4F8 | 4.096 |
| 25 | EIF2AK2 | P19525 | 25.60 |
| 26 | CLSTN3 | Q9BQT9 | 1.638 |
| 27 | GLRX | P35754 | 0.655 |
| 28 | NLGN1 | Q8N2Q7 | 0.262 |
| 29 | SYT1 | P21579 | 0.042 |
| 30 | DDAH1 | O94760 | 0.655 |
| 31 | SNCAIP | Q9Y6H5 | 0.007 |
| 32 | BACE1 | P56817 | 1.638 |
| 33 | LRRK2 | Q5S007 | 1.638 |
| 34 | AGER | Q15109 | 4.096 |
| 35 | WWOX | Q9NZC7 | 0.017 |
| 36 | NPTX2 | P47972 | 25.60 |
| 37 | ENO2 | P09104 | 4.096 |
| 38 | KLK8 | O60259 | 1.638 |
| 39 | SCG2 | P13521 | 4.096 |
| 40 | TP53 | P04637 | 0.003 |
| 41 | RTN4R | Q9BZR6 | 0.655 |
| 42 | MOG | Q16653 | 0.042 |

Supplementary Table 2. Assay comparisons between methods

| Assay | Platform | Matrix |
|---|---|---|
| Abeta42 | NULISA, Elecsys, Simoa | CSF and plasma |
| Abeta40 | NULISA, Simoa | CSF and plasma |
| NfL | NULISA, Simoa | CSF and plasma |
| GFAP | NULISA, Simoa | CSF and plasma |
| BACE1 | NULISA | CSF and plasma |
| ENO2 | NULISA | CSF and plasma |
| NGF | NULISA | Plasma |
| NPTX1 | NULISA | CSF and plasma |
| NPTX2 | NULISA | CSF and plasma |
| NPTXR | NULISA | Plasma |
| pTau217 | NULISA | CSF and plasma |
| S100B | NULISA | CSF |
| SMOC2 | NULISA | CSF and plasma |
| sTREM1 | NULISA | CSF and plasma |
| sTREM2 | NULISA | CSF and plasma |
| VSNL1 | NULISA | CSF and plasma |

Supplementary Table 3. Individual assay CV%

| **Assay** | **Intra-plate CV%** | **Inter-plate CV%** |
|---|---|---|
| AGER | 5.33 | 9.91 |
| Abeta40 | 5.36 | 4.90 |
| Abeta42 | 8.09 | 8.40 |
| BACE1 | 4.30 | 6.74 |
| BMP7 | 6.67 | 6.78 |
| CLSTN3 | 5.75 | 6.38 |
| DDAH1 | 1.11 | 13.02 |
| DDC | 5.07 | 7.45 |
| EIF2AK2 | 9.18 | 6.94 |
| ENO2 | 8.06 | 8.76 |
| FOXO3 | 6.14 | 5.89 |
| GDNF | 3.73 | 3.88 |
| GFAP | 7.13 | 8.90 |
| GLRX | 5.39 | 7.88 |
| HLA-DRA | 4.25 | 6.30 |
| ITGAM | 4.46 | 7.35 |
| ITGB2 | 6.65 | 5.86 |
| KLK8 | 7.00 | 9.58 |
| LRRK2 | 5.83 | 8.18 |

| | | |
|---|---|---|
| MMP10 | 7.20 | 8.07 |
| MOG | 7.69 | 9.87 |
| NfL | 4.16 | 4.75 |
| NGF | 7.98 | 6.72 |
| NLGN1 | 6.25 | 6.96 |
| NPTX1 | 5.79 | 7.11 |
| NPTX2 | 6.93 | 9.74 |
| NPTXR | 8.85 | 9.00 |
| OMG | 6.81 | 8.85 |
| RTN4R | 4.74 | 8.39 |
| S100B | 8.34 | 9.24 |
| SCG2 | 5.93 | 9.26 |
| SDC4 | 4.30 | 5.80 |
| SMOC1 | 8.27 | 9.39 |
| SNCAIP | 4.44 | 2.93 |
| STX1B | 7.89 | 11.34 |
| SYT1 | 9.52 | 10.64 |
| TP53 | 5.60 | 5.88 |
| sTREM1 | 7.32 | 7.25 |
| sTREM2 | 7.85 | 7.33 |
| VSNL1 | 11.50 | 7.44 |
| WWOX | 4.76 | 5.34 |
| pTau217 | 5.64 | 10.00 |

Coefficient of variation percentage (CV%) were calculated based on the sample controls included in each plate. All assays show a CV below 15%. The median intra-plate and inter-plate CVs for each individual assay are shown. In addition, the overall median intra-plate CV% and the intra-plate CV% were calculated as 7.4% and 6.5%, respectively.

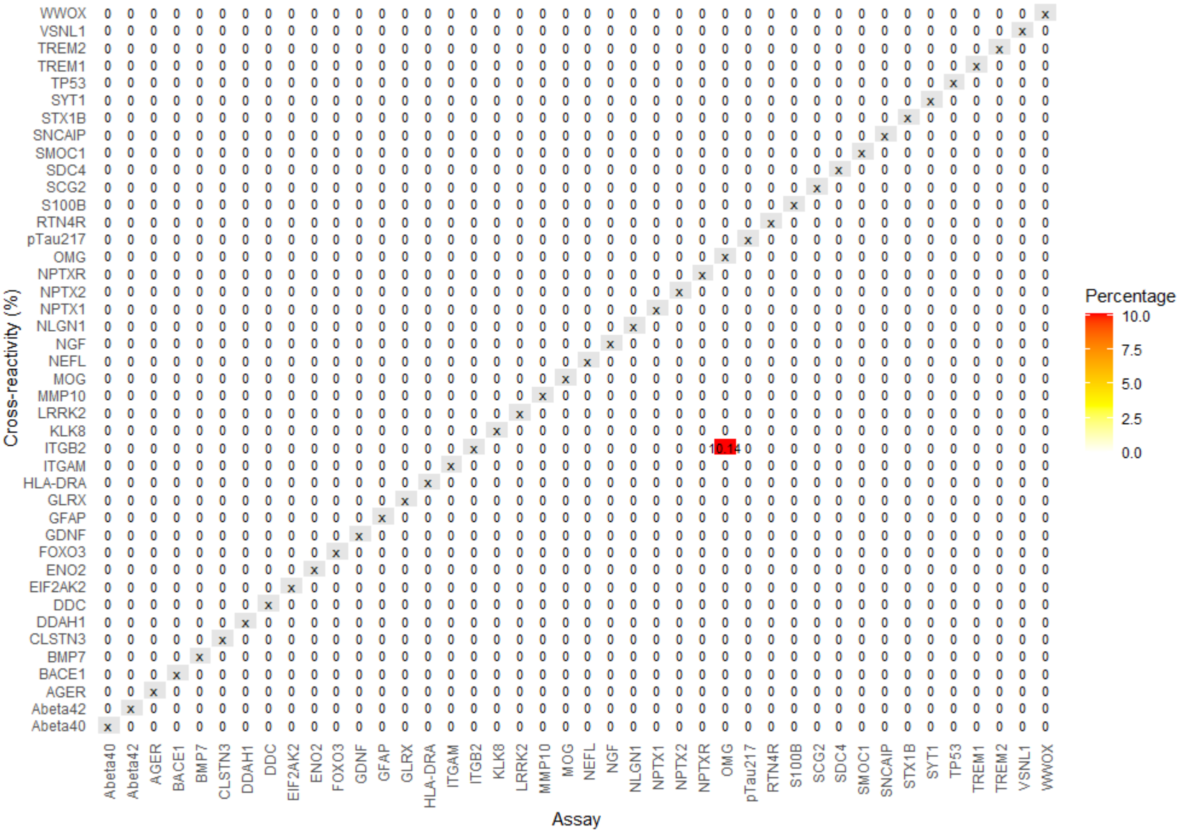


### Supplementary Figure 1. Specificity test

Recombinant antigens for all 42 assays were measured in duplicate, with a pooled antigen mixture as positive control. Target concentrations were based on EDTA plasma signal intensities, and non-specific signal was expressed relative to true signal. Cross-reactivity was detected for the OMG assay specifically, with a 10% signal increase observed upon addition of the ITGB2 antigen.

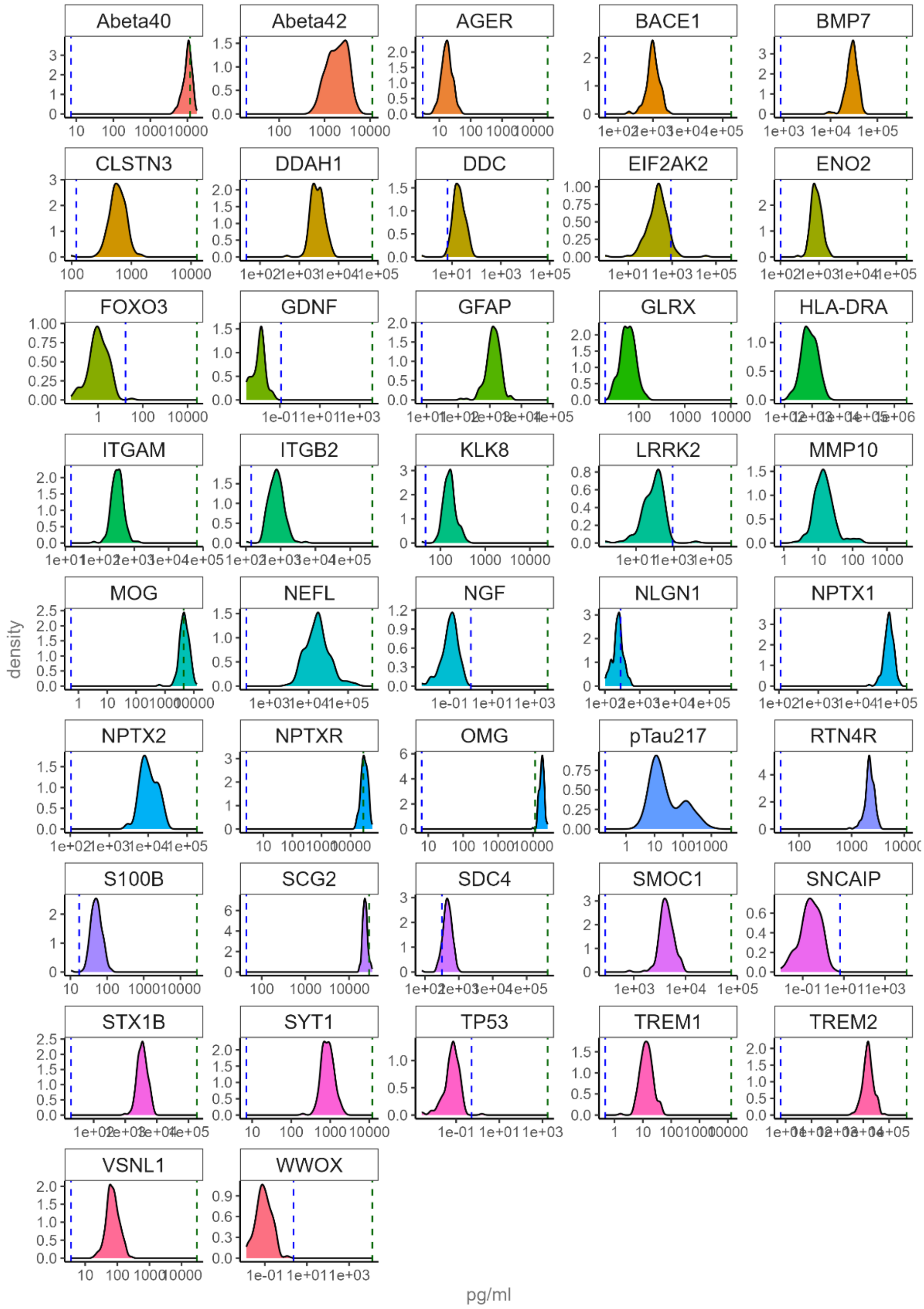


Supplementary Figure 2. Sample detectability in CSF

Density plots display the distribution of protein concentrations (pg/mL) of the CSF samples above the lower limit of quantification (LLOQ, dotted vertical blue line), within the quantifiable range, and below the upper limit of quantification (ULOQ, dotted vertical green line) for each assay. Eight assays had results below the LLOQ (i.e., LRRK2, GDNF, WWOX, FOXO3, NGF, EIF2AK2, TP53, and SNCAIP) and three assays had results above the ULLOQ (i.e., OMG, MOG, and NPTXR).

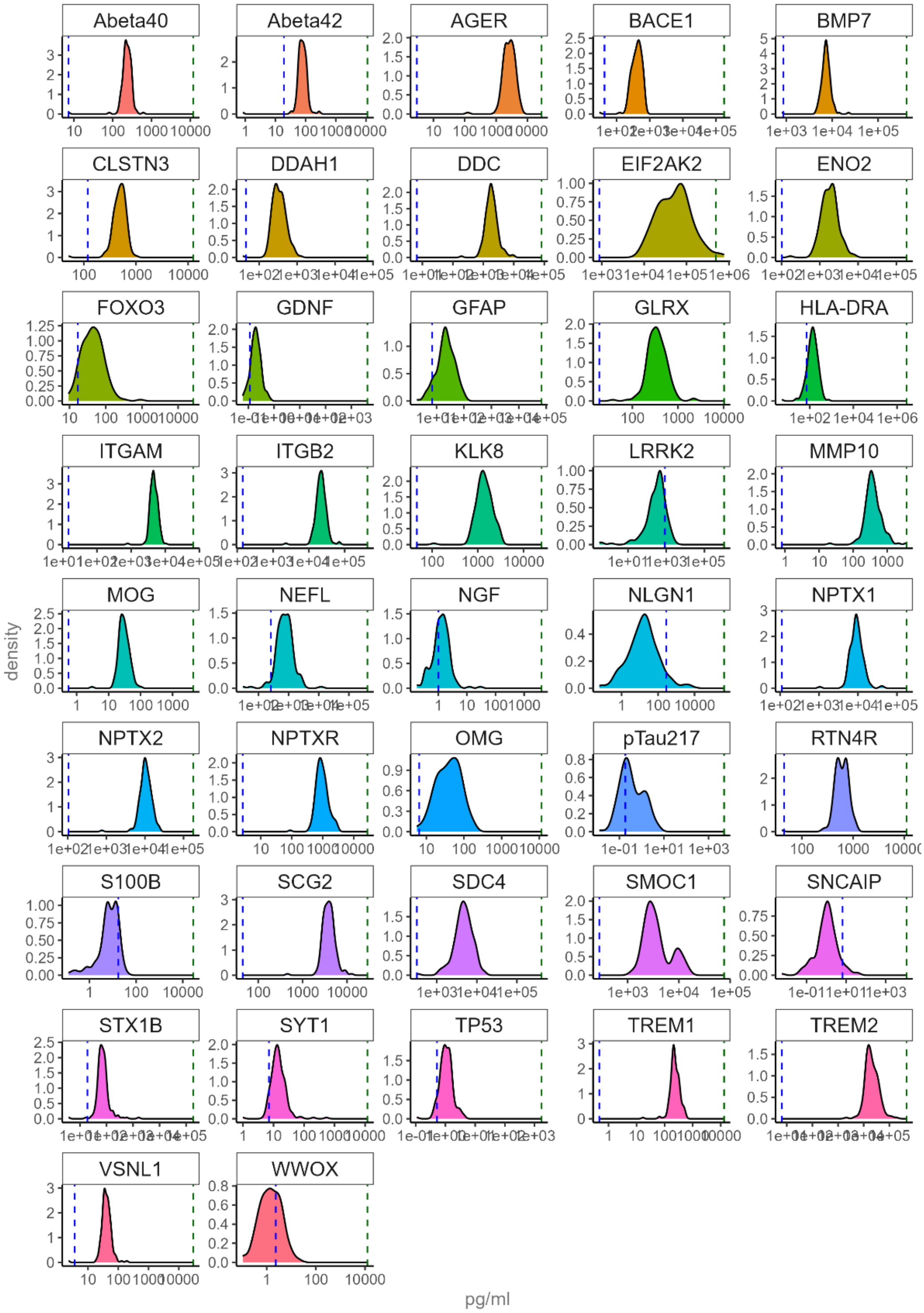


Supplementary Figure 3. Sample detectability in plasma

Density plots display the distribution of protein concentrations (pg/mL) of the plasma samples above the lower limit of quantification (LLOQ, dotted vertical blue line), within the quantifiable range, and below the upper limit of quantification (ULOQ, dotted vertical green line) for each assay. Results were below the LLOQ in four assays (i.e., S100B, LRRK2, NLGN1, and SNCAIP).

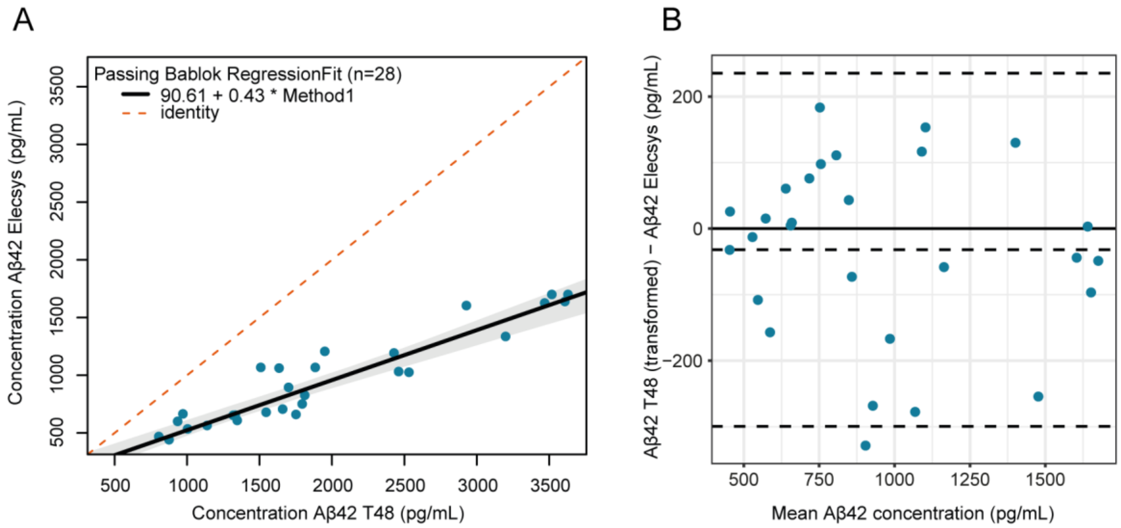


Supplementary Figure 4. Platform comparison with Elecsys using CSF samples

A) Passing-Bablok regression of Aβ42 concentrations measured by the T48 Neuropanel and Elecsys assays (slope: 0.43, 95% CI: 0.37 – 0.48 and intercept: 90.61, 95% CI: 6.16 – 205.03). The solid line represents the regression line, the grey area shows 95% CI of the regression line, and the dashed line represents the identity line (x = y). Method 1 is defined as the T48 Neuropanel. B) Bland-Altman plot shows the difference in Aβ42 concentrations (pg/mL) between the T48 Neuropanel and Elecsys assays after transformation of the Aβ42 T48 values to Elecsys (T48 Neuropanel minus Elecsys). The solid grey line represents the bias (-32.04 pg/mL, 95% CI: -84.93 – 20.86 pg/mL), dashed lines are the upper (235 pg/mL) and lower (-299.42 pg/mL) 95% limits of agreement.

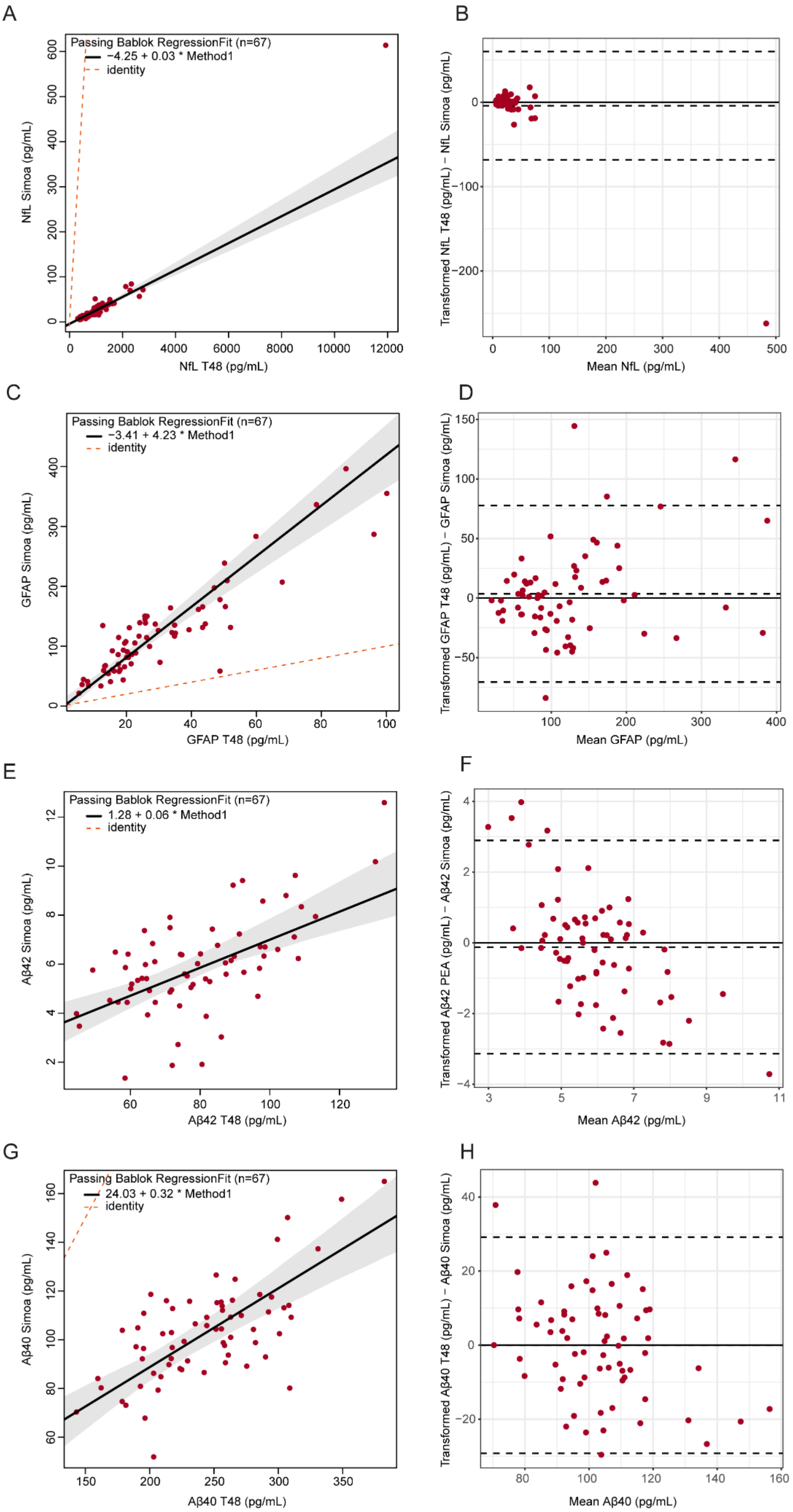


Supplementary Figure 5. Platform comparison with Simoa using plasma samples

Passing-Bablok regression formulas for NfL (A; slope: 0.03, 95% CI: 0.0259 – 0.0353 and intercept: -4.25, 95% CI: -8.42 – -0.99), GFAP (C; slope: 4.23, 95% CI: 3.59 – 4.77 and intercept: -3.41, 95% CI: -15.11 – 10.01), Aβ42 (E; slope: 0.06, 95% CI: 0.04 – 0.08 and intercept: 1.28, 95% CI: -0.24 – 2.88), and Aβ40 (G; slope: 0.32, 95% CI: 0.24 – 0.42 and intercept: 24.03, 95% CI: -0.09 – 44.49) assays measured by the T48 Neuropanel and Simoa. The solid line represents the regression line, the grey area shows 95% CI of the regression line, and the dashed line represents the identity line (x = y). Method 1 is defined as the T48 Neuropanel. Bland Altman plots showed small non-significant bias for NfL (B; bias = -3.91 pg/mL, 95% CI: -11.85 – 4.04, upper limit = 59.92 pg/mL, lower limit = -67.73), GFAP (D; bias = 3.59 pg/mL, 95%CI: -5.64 – 12.82, upper limit = 77.75 pg/mL, lower limit = -70.57), Aβ42 (F; bias = 0.10 pg/mL, 95% CI: -0.3 – 0.47, upper limit = 3.11 pg/mL, lower limit = -2.91), and Aβ40 (H; bias =-0.89 pg/mL, 95% CI: -4.52 – 2.73, upper limit = 28.26 pg/mL, lower limit = -30.05) after transformation of the T48 values to the Simoa scale using the Passing-Bablok transformation formulas.

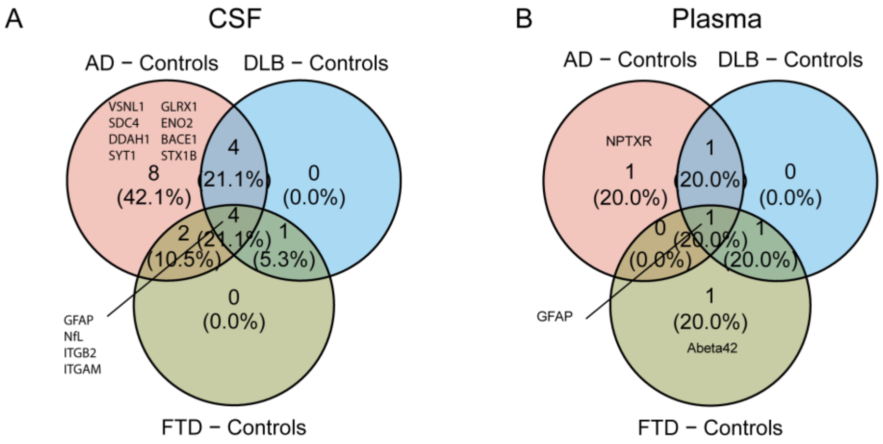


Supplementary Figure 6. Overlapping protein changes in CSF and plasma across AD, DLB, and FTD dementias.

Venn diagrams show FDR-corrected significantly dysregulated proteins in CSF (A) and plasma (B). In CSF, four proteins were significantly altered across all AD, FTD, and DLB groups (GFAP, NfL, ITGB2, and ITGAM), while in plasma, only GFAP was significantly altered across groups.
